# Systematic Review and Meta-Analysis on the Prevalence of *Taenia solium* Infections Across India

**DOI:** 10.1101/2025.06.23.25330151

**Authors:** Rashmi Sharma, R.S. Aulakh, Rajnish Sharma, Balbir Singh Bagicha

## Abstract

**Background:** *Taenia solium* taeniasis and cysticercosis continue to pose significant public health and veterinary challenges in endemic regions like India. Despite sporadic reports, a comprehensive understanding of the national burden has been lacking. This systematic review and metaanalysis aimed to estimate the pooled prevalence of *T. solium* infection in humans and pigs across India and assess regional, demographic, and methodological variations.

**Methods:** A systematic search of six international databases (PubMed, Scopus, Medline, Embase, Web of Science, Google Scholar) and national repositories (Shodhganga, Krishikosh, CeRA) was conducted for studies published between 1950 and December 2021. Studies reporting prevalence of human or porcine taeniasis and/or cysticercosis in India were included. Data were extracted from 67 eligible studies (45 human, 22 porcine) and analyzed using random effects models to account for heterogeneity. Subgroup analyses examined variations by zone, setting, gender, and diagnostic modality.

**Results:** The pooled prevalence of *T. solium* infection in humans was 5.21% (95% CI: 5.00–5.43%), based on data from 41,481 individuals. Prevalence varied significantly across regions, with the highest in the North-East Zone (12.00%) and South Zone (8.08%). Hospital-based studies reported substantially higher prevalence (26.18%) compared to population-based studies (4.64%), likely reflecting diagnostic bias toward symptomatic cases. Cysticercosis prevalence (10.24%) exceeded that of taeniasis (3.69%). Female participants showed higher prevalence (4.26%) than males (3.08%) (p < 0.0001). In pigs, the overall pooled prevalence was 3.27%, with higher burdens observed in the Central and East/North-East zones.

**Conclusions:** This meta-analysis confirms that *T. solium* infection remains endemic in India, with marked heterogeneity across zones and study settings. The strong human–porcine epidemiological link highlights the need for a tailored One Health strategy incorporating regional risk factors, public health education, improved diagnostics, and sanitation infrastructure. Enhanced surveillance in high-prevalence zones like the North-East and South is urgently needed to mitigate transmission.

## Introduction

*Taenia solium* taeniasis/cysticercosis is a parasitic zoonosis prioritized by the World Health Organization as a neglected tropical disease that poses a significant threat to public health and economic stability in developing nations (WHO, 2023). The parasite’s complex life cycle involves humans as the sole definitive hosts for the adult tapeworm (taeniasis) and both pigs and humans as intermediate hosts for the larval stage (cysticercosis) (Garcia *et al*., 2003). When these larvae encyst in the central nervous system, the resulting condition, neurocysticercosis (NCC), is a major cause of acquired epilepsy worldwide and is considered the leading cause of preventable epilepsy in endemic areas (Garcia & Del Brutto, 2005; Ndimubanzi *et al*., 2010). The burden of this disease is not only measured in human suffering but also in significant economic losses due to healthcare costs and reduced productivity in both human and agricultural sectors (Rajshekhar *et al*., 2003).

In India, a combination of socioeconomic and environmental factors creates conditions conducive to the parasite’s transmission. These include inadequate sanitation infrastructure and the widespread practice of open defecation, which facilitates environmental contamination with *T. solium* eggs (Dixon *et al*., 2019). Furthermore, the prevalence of traditional, free-roaming pig husbandry systems, particularly in rural and peri-urban communities, allows pigs to easily encounter human feces, thus perpetuating the parasite’s life cycle (Thomas *et al*., 2013). This “One Health” paradigm, where human health, animal health, and the environment are inextricably linked, is central to understanding and controlling *T. solium* (Zinsstag *et al*., 2011).

While numerous epidemiological studies have been conducted across India, they have produced a wide range of prevalence estimates (Singh *et al*., 2014). This variability, often dependent on the specific geographical location, study design, and diagnostic methods used, makes it challenging to ascertain the true national burden of the disease and to formulate effective, evidence-based control strategies (Prasad *et al*., 2008). A comprehensive, updated synthesis of the available data is therefore crucial for public health planning.

The primary objective of this study was to conduct a systematic review and meta-analysis of published literature to estimate the overall pooled prevalence of *T. solium* taeniasis and cysticercosis in both human and porcine populations in India. Additionally, this study aimed to investigate the geographical distribution of the parasite across different administrative zones and to identify key demographic risk factors associated with infection.

### Methodology

#### Search Strategy

A systematic and comprehensive literature search was executed to identify all relevant studies concerning the prevalence of *Taenia solium* taeniasis and cysticercosis within India. The geographical scope of this review was exhaustive, encompassing all 28 states and 8 union territories. The search period covered publications from 1950 up to and including December 2021. The search strategy was designed to be highly sensitive and included multiple major international electronic databases: PubMed, Scopus, Medline, Embase, Web of Science, and Google Scholar. This was done to ensure a broad capture of peer-reviewed literature.

A specific Boolean search query was constructed and applied consistently across these platforms, utilizing the following keywords and operators: (“Prevalence” OR “epidemiology”) AND (“*Taenia solium* “ OR “cysticercosis” OR “neurocysticercosis” OR “porcine cysticercosis” OR “taeniasis”) AND (“India”). To ensure a comprehensive review of Indian-specific research, the search was also extended to national academic and agricultural repositories, including Shodhganga, Krishikosh, and the Consortium for e-Resources in Agriculture (CeRA).

To identify studies not indexed in these electronic databases, a manual search for “grey literature” was conducted. This involved a thorough review of materials such as academic theses, dissertations, institutional annual reports, and conference proceedings obtained from various university libraries and the specialized library of the Department of Veterinary Public Health and Epidemiology. Finally, the reference lists of all articles selected for inclusion were meticulously hand-searched to identify any additional studies that may have been missed during the initial database searches.

#### Study Screening and Selection

The retrieved articles were subjected to a rigorous, multi-stage screening process to determine their eligibility for inclusion in the final meta-analysis. In the first stage, the titles and abstracts of all collected articles were independently screened for relevance. During this initial phase, publications were categorized as either potentially relevant or irrelevant based on the presence of key terms such as “prevalence,” “*Taenia solium*,” “porcine cysticercosis,” and “taeniasis.”

Studies that passed this preliminary screening advanced to the second stage: a full-text eligibility assessment. Each article was carefully reviewed against a predefined set of inclusion criteria. To be included, a study had to: (1) be conducted within India; (2) be published in the English language; (3) report on primary research findings; (4) be a population-based or a hospital based study; (5) involve either human or porcine populations; and (6) provide precise, extractable data on the study’s sample size, the number of positive cases identified, and the distribution of associated risk factors.

Studies that did not me*et* all these criteria were excluded. After systematically removing duplicate records from the pool of eligible articles, a total of 882 unique studies were identified as relevant. Following the detailed full-text review, 67 of these studies were confirmed to me*et al*l inclusion criteria and were carried forward for data extraction and inclusion in the meta-analysis. The entire selection process is transparently documented in a PRISMA flow diagram (Fig. 1).

**Fig. 1:**
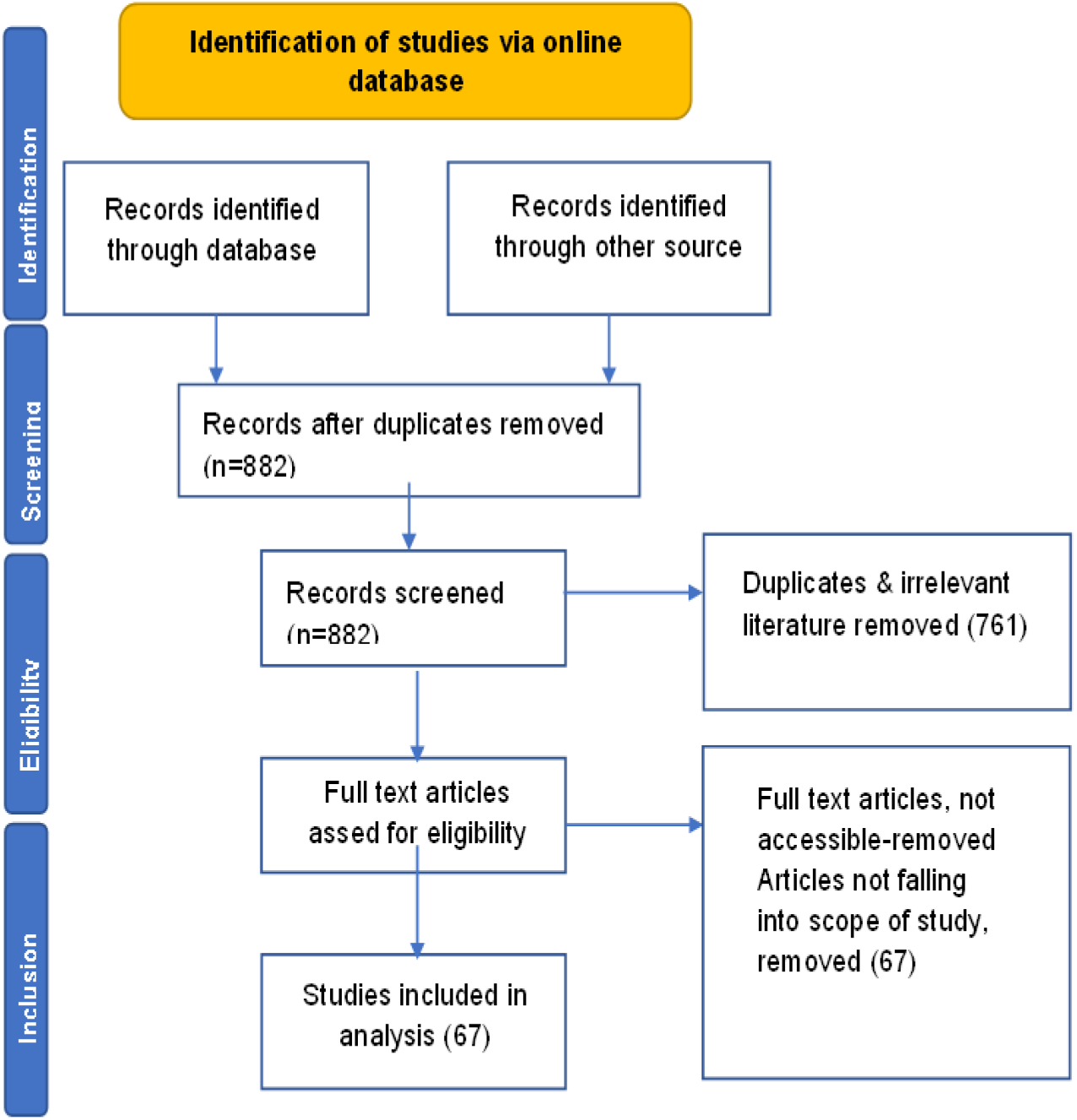
Flowchart of literature collection as PRISMA Guidelines

#### Data Extraction

A standardized data extraction form was developed using a Microsoft Excel spreadsheet (2019) to ensure consistency and accuracy in data collection. Information from each of the 67 selected studies was carefully extracted and entered to a spreadsheet. The key data points recorded for each study included the article title, the first author’s name, the year of publication, and the journal of publication. A comprehensive list outlining all specific parameters that were extracted for the purpose of the meta-analysis is provided in Table 1.

**Table 1:**
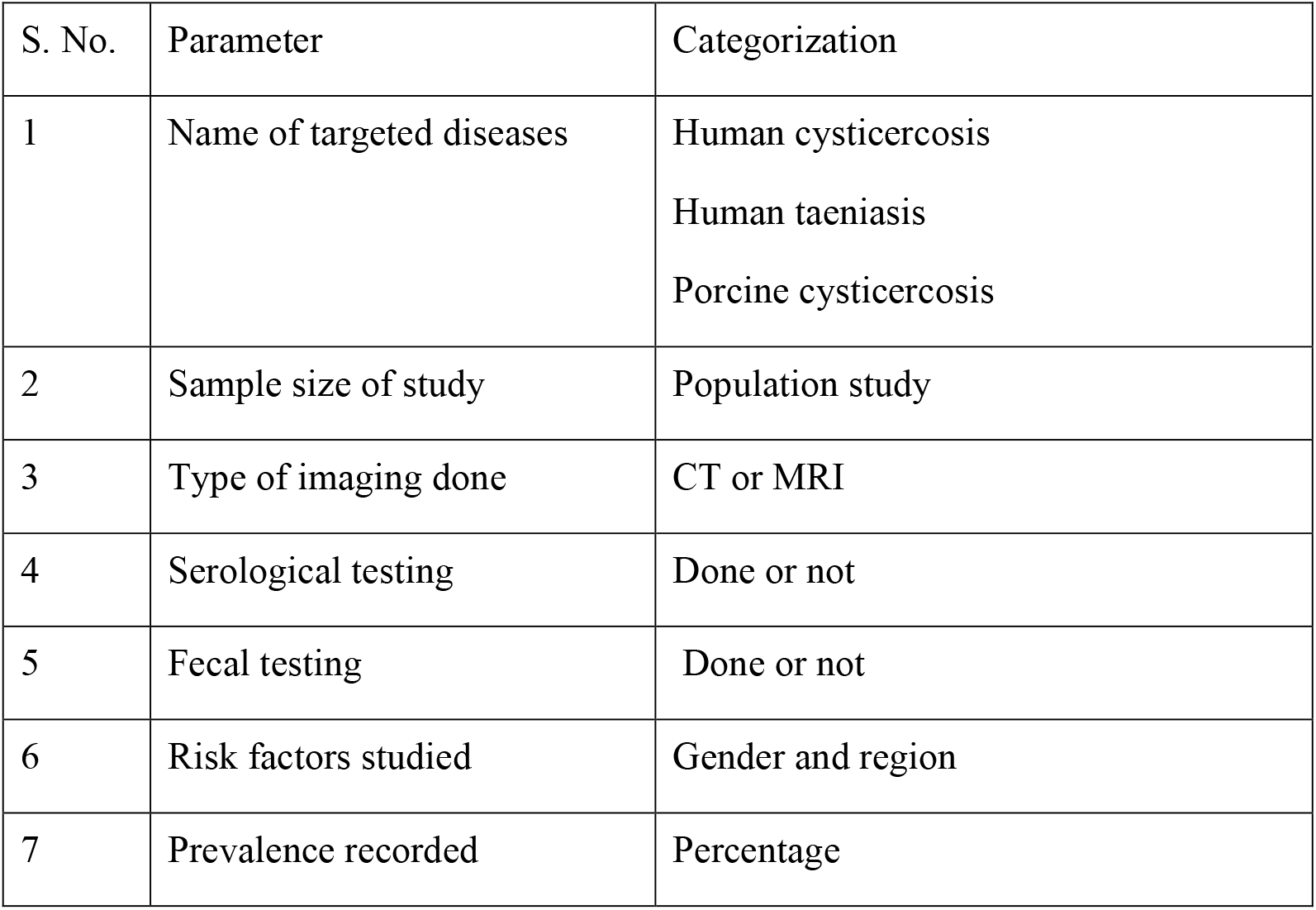
Data Parameters Extracted for Meta-Analysis and Systematic Review.

## Results

Following a comprehensive search and screening process that initially identified 882 unique articles, a final selection of 67 studies met the stringent inclusion criteria for this meta-analysis. The included research represents a broad investigation into *Taenia solium* across India. A substantial portion of the research, 45 studies, was dedicated to investigating human populations, while the remaining 22 studies focused specifically on porcine populations. This body of evidence allowed for a synthesized analysis of the prevalence of human taeniasis, the various manifestations of human cysticercosis (including neurocysticercosis and ocular cysticercosis), and porcine cysticercosis.

The analysis of the 45 studies focused on human health incorporated data from a large sample of 41,481 individuals from diverse communities across India. The resulting data revealed a significant overall pooled prevalence of *T. solium* infection (which includes both taeniasis and cysticercosis) estimated at 5.21% (95% CI: 5.00–5.23%), indicating that the parasite remains a notable public health concern in the country.

The burden of *T. solium* infection is not uniformly distributed across India, with prevalence rates varying markedly among the nation’s six administrative zones. The North Zone was the most frequently studied region (n=18), yielding a pooled prevalence of 3.29% (95% CI: 3.07–3.53%) from a sample of 22,741 individuals. In stark contrast, the highest prevalence was recorded in the North-East Zone at 12.00% (95% CI: 7.73–18.17%), a finding derived from a single study of 150 individuals. The South Zone, another well-represented region with 17 studies, demonstrated a high prevalence of 8.08% (95% CI: 7.59–8.59%; 930/11,516). Other zones showed varied prevalence rates: the West Zone (n=2 studies) at 7.59% (95% CI: 6.25–9.14%), the Central Zone (n=7 studies) at 3.26% (95% CI: 2.99–3.55%), and the East Zone (n=1 study) at 4.48%. The geographical disparities in prevalence are visually summarized in the map presented in Fig. 2.

**Figure 2.**
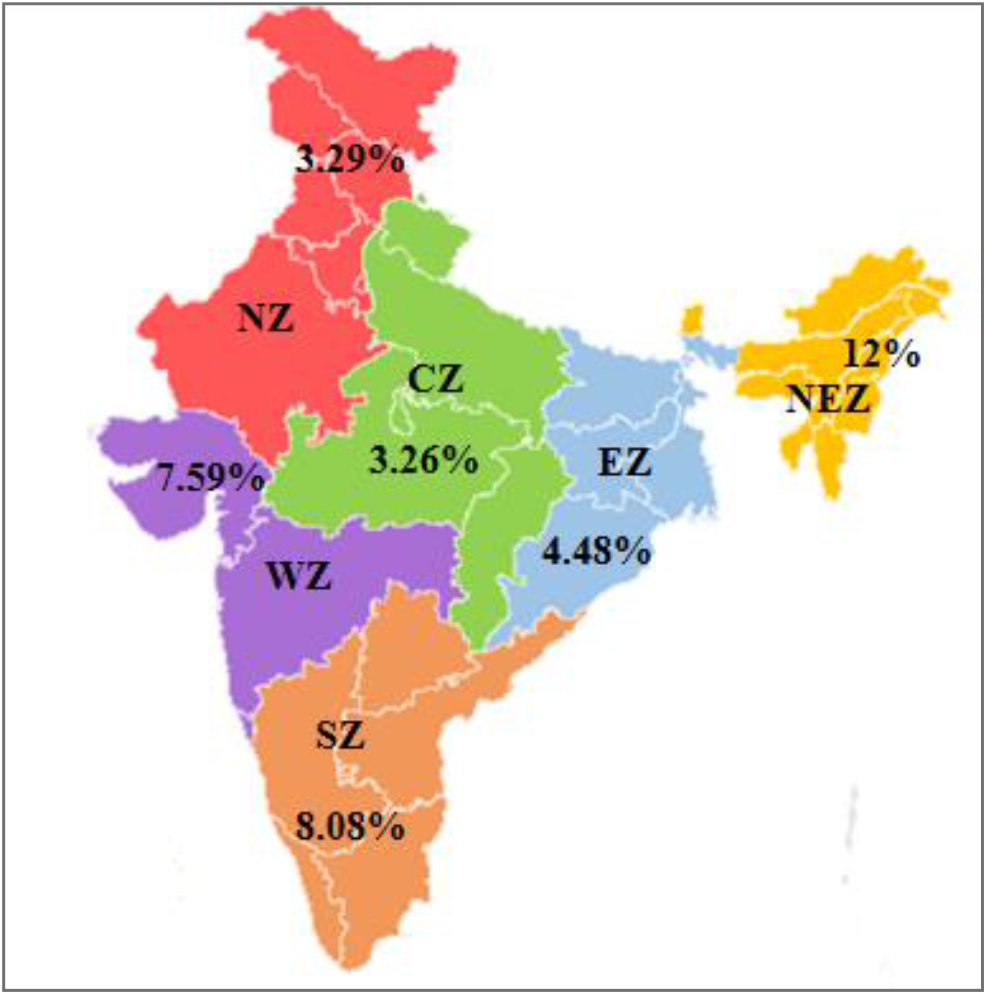
Map showing Zone wise Prevalence rate in of *Taenia solium* in India

A critical distinction emerged when analyzing prevalence based on the study setting. The 32 population-based studies, reflecting community-level exposure, indicated a pooled prevalence of 4.64% (95% CI: 4.44–4.85%). Conversely, the 13 hospital-based studies, which predominantly focused on patients presenting with neurological symptoms, revealed a much higher apparent pooled prevalence of 26.18% (95% CI: 24.44–28.00%). This inflation is expected, as these clinical settings concentrate cases of neurocysticercosis, for which computed tomography (CT) and magnetic resonance imaging (MRI) were the primary diagnostic tools.

When the infections were differentiated by type, the analysis showed an overall taeniasis prevalence of 3.69% (95% CI: 3.44–3.95%) in the general population. The prevalence of human cysticercosis was notably higher at 10.24% (95% CI: 9.76–10.44%), highlighting the substantial burden of the larval stage of the parasite.

### Association with Gender

The analysis identified gender as a significant risk factor for *T. solium* infection in humans. A higher prevalence was observed among females (4.26%) compared to males (3.08%), as illustrated in Fig. 3. This association was found to be statistically significant (chi-square test, p < 0.0001), suggesting that females may be at greater risk of exposure.

**Figure. 3:**
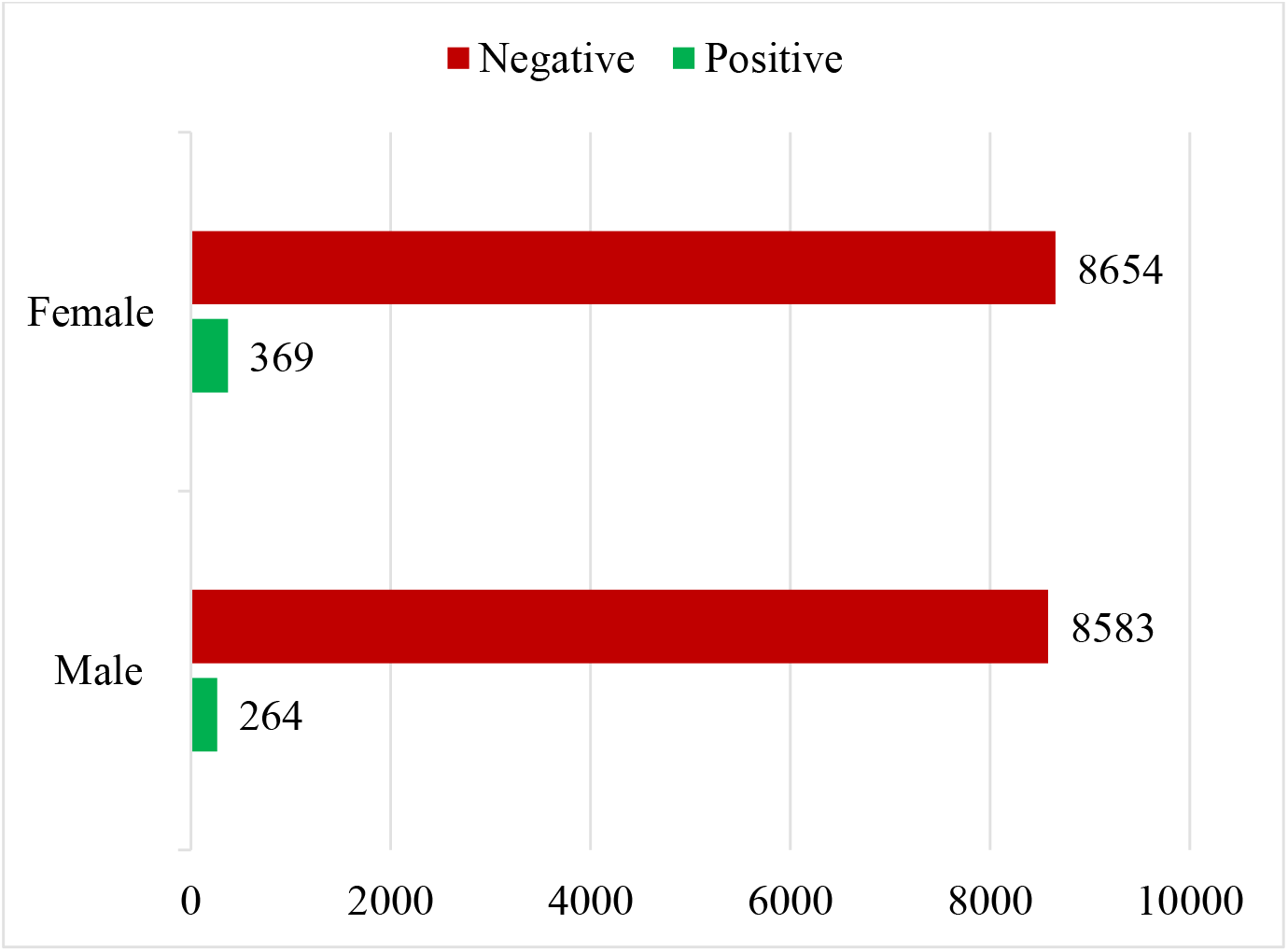
Number of males and females found positive for *Taenia solium* infection

### Forest Plot Analysis of Human Infection

The meta-analysis of 45 studies on human infection showed extreme heterogeneity (I^2^ = 99.8%, p < 0.001). This indicates that the substantial variation in prevalence rates across the studies is due to true underlying differences in factors like geography, study design (hospital-vs. population-based), and diagnostic methods, rather than random sampling error. The reported prevalence in individual studies demonstrated a vast range, from 0% in some community surveys to as high as 100% in hospital-based studies focusing on patients with epilepsy. This wide dispersion visually represents the heterogeneity captured by the I^2^ statistic.

Due to this high level of heterogeneity, a random-effects model was used to calculate the pooled estimate. This model yielded an overall pooled prevalence of 5.21% (95% CI: 5.00%–5.43%). This estimate provides a statistical average of human *T. solium* infection across the included studies, accounting for the wide variability. The forest plot analysis reinforces the conclusion that prevalence is highly dependent on the population being studied, with clinical settings showing a much higher burden of disease (Figure 4).

**Figure 4.**
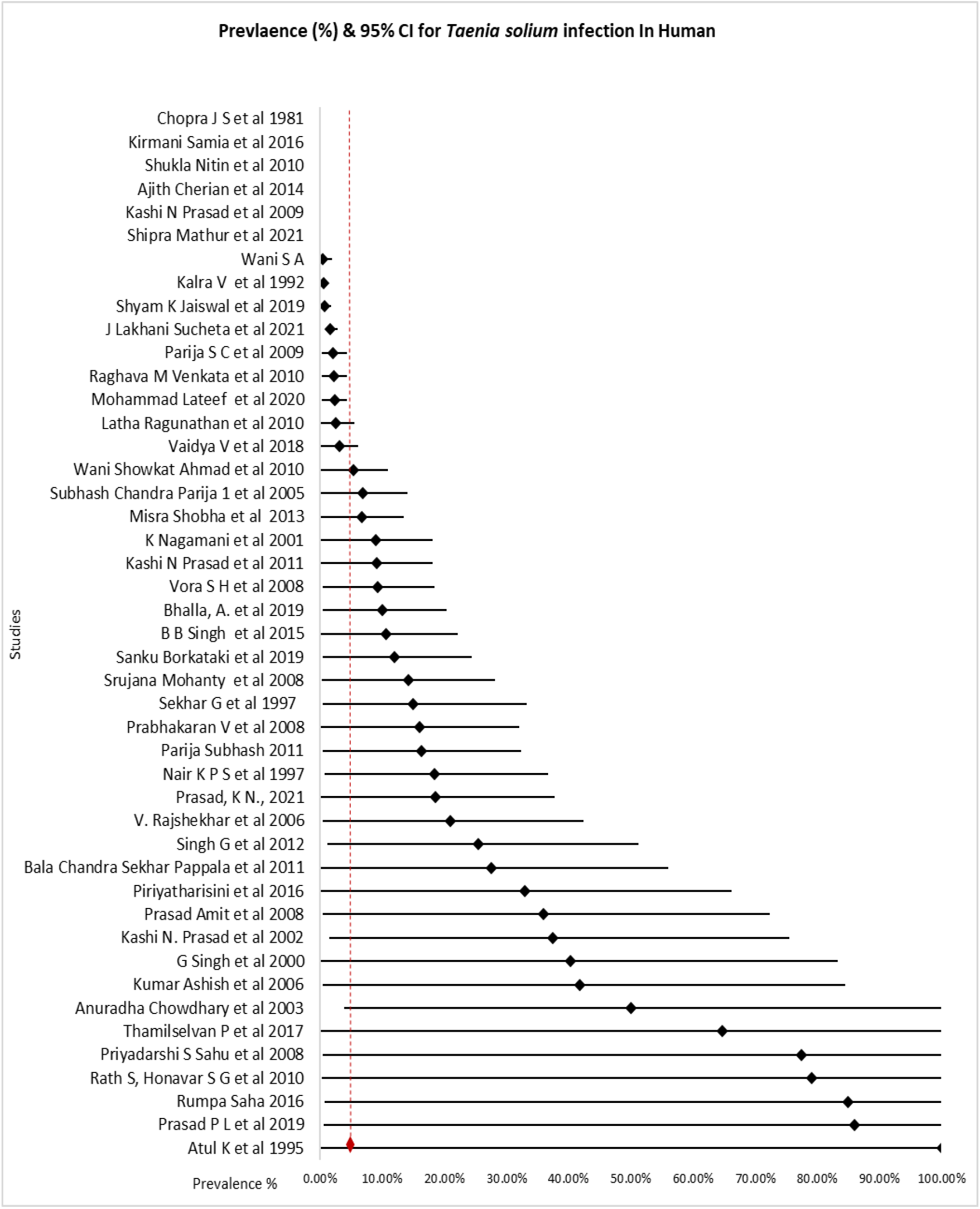
Forest Plot of *Taenia solium* Infection Prevalence (95% CI) Across 45 Studies in India

### Burden Of Porcine Cysticercosis in India

The overall prevalence of porcine cysticercosis was 3.27%. The geographical distribution of porcine cysticercosis mirrored the human data to some extent, with high prevalence in the Central (7.60%) and East/North-East (7.30%) zones. In contrast to humans, there was no gender-based difference in prevalence among pigs.

### Forest Plot Analysis of Porcine Cysticercosis

The meta-analysis of 22 studies on porcine cysticercosis revealed extreme heterogeneity among the studies (I^2^ = 99.4%, p < 0.001). This statistically significant level of heterogeneity indicates that the observed variation in prevalence is not due to random chance but reflects true differences in infection rates across the diverse geographical regions and study populations included in the analysis. The prevalence reported in individual studies ranged widely, from as low as 0.3% to as high as 20.75%, visually confirming this substantial variability.

Given the high heterogeneity, a random-effects model was employed to determine the overall pooled prevalence. This model yielded a pooled prevalence of 3.27%. This estimate, which accounts for the variation between studies, provides a robust summary of the average prevalence of porcine cysticercosis across India (Figure 5).

**Figure 5.**
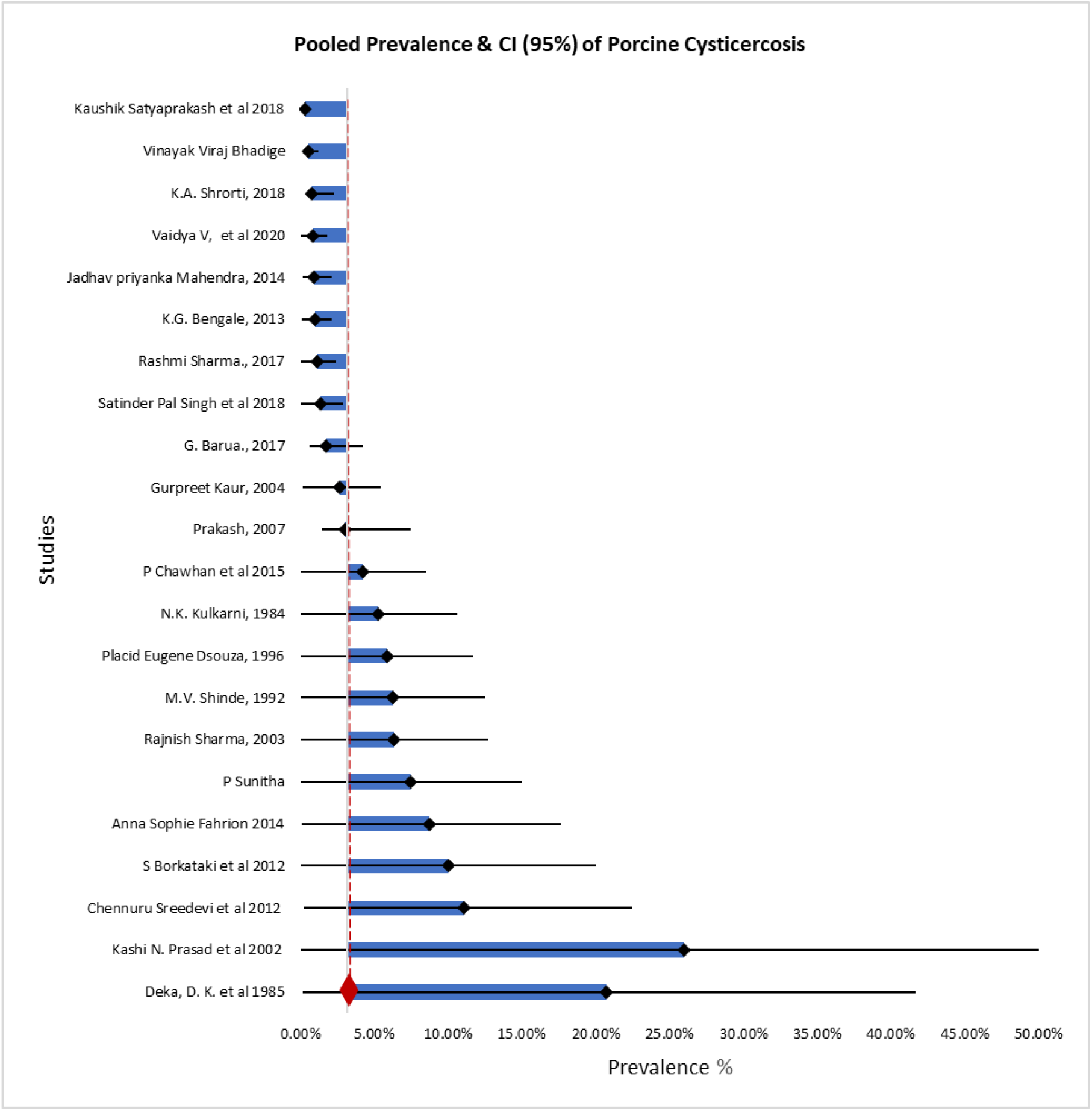
Forest Plot Prevalence of Porcine cysticercosis from 22 studies in India

## Discussion

This systematic review and meta-analysis provide a comprehensive update on the prevalence and distribution of *Taenia solium* taeniasis and cysticercosis in both human and porcine populations across India. The findings confirm that *T. solium* infection is endemic and represents a significant, ongoing public health and veterinary challenge in the country. The overall pooled prevalence in humans was 5.21%, and in pigs was 3.25%, highlighting the persistent circulation of the parasite.

The estimated human prevalence of 5.21% in our study is a cause for concern. When compared to a previous meta-analysis focused on the broader Asian continent, which reported a prevalence of 3.89% (Bizhani *et al*., 2021), our findings suggest that the burden in India may be comparatively higher. However, our result is lower than the 12.7% seroprevalence reported in a review of studies from Latin America (Coral-Almeida *et al*., 2015), where the disease is famously hyperendemic. A study in neighboring Nepal reported a community-based seroprevalence of neurocysticercosis at 8.2%, which is comparable to the prevalence we found in India’s South Zone (Sako *et al*., 2020).

A key finding of our analysis is the stark discrepancy between hospital-based (26.18%) and population-based (4.64%) prevalence. This vast difference is consistent with findings from other endemic regions (Ndimubanzi *et al*., 2010; O’Neal *et al*., 2012) and underscores the nature of hospital data, which is skewed towards symptomatic cases, particularly neurocysticercosis— the most severe form of the disease. While population-based studies provide a more accurate reflection of community-level exposure, the high prevalence in hospital settings highlights the severe clinical burden and economic impact of the disease (Rajshekhar *et al*., 2003; Singh *et al*., 2014).

Our analysis revealed significant geographical heterogeneity in human prevalence across India’s administrative zones. The extremely high prevalence in the North-East Zone (12.00%), though based on a single study, aligns with reports from other researchers who have identified this region as a hotspot due to specific socio-cultural factors, including traditional pig-rearing practices and dietary habits (Roy *et al*., 2018; Devi *et al*., 2021). The high prevalence in the South Zone (8.08%) is also well-documented, with numerous studies from states like Tamil Nadu and Andhra Pradesh consistently reporting a high disease burden (Prasad *et al*., 2007; Murrell, 2013). The lower prevalence in the North Zone (3.29%) is surprising given the large number of studies, but this may reflect diverse local conditions within a very large geographical area. This zonal variation emphasizes that control strategies must be tailored to local epidemiological contexts rather than a one-size-fits-all national policy (Li *et al*., 2019).

Another significant finding was the higher prevalence in females (4.26%) compared to males (3.08%). This contrasts with some studies that found no gender difference (Fleury *et al*., 2006) but is consistent with others that have suggested females are at higher risk (Kumar *et al*., 2011; Panicker *et al*., 2012). This increased risk may be attributed to greater involvement in food preparation and potential exposure to contaminated vegetables or water within the household setting (Garcia *et al*., 2003; Montresor *et al*., 2017).

The overall prevalence of porcine cysticercosis was 3.27%. This figure is lower than the 5.21% reported in a recent meta-analysis by Bhangale *et al*. (2021). As noted in our results, this discrepancy can be partially attributed to methodological differences; our study primarily included data from postmortem examinations, providing definitive evidence of infection, whereas the analysis by Bhangale and colleagues also included serological studies, which can indicate past exposure rather than active infection and may lead to higher estimates (Dorny *et al*., 2003). Our prevalence is comparable to rates reported in other Southeast Asian countries like Vietnam and Cambodia (Van De *et al*., 2016; Ty *et al*., 2017) but lower than rates in parts of sub-Saharan Africa, where prevalences can exceed 20% (Krecek *et al*., 2008; Thomas *et al*., 2015).

The geographical distribution of porcine cysticercosis mirrored the human data to some extent, with high prevalence in the Central (7.60%) and East/North-East (7.30%) zones. This spatial correlation is expected and reinforces the link between pig husbandry practices, environmental contamination, and human infection risk (Lightowlers *et al*., 2007; Praet *et al*., 2010). The low prevalence in the West Zone (1.50%) despite many studies may reflect more developed and controlled pig farming systems in that region.

Unlike human studies, prevalence rate was not affected by gender of animal consistent with the various previous studies and existing literature (Phiri *et al*., 2003; Ganaba *et al*., 2011). As exposure for free-roaming pigs is largely determined by their access to human feces in the environment, which is independent of sex.

## Limitations and Future Directions

This meta-analysis has several limitations. The reliance on a small number of studies in certain zones (e.g., North-East and East) necessitates caution in interpreting the zonal prevalence. Furthermore, the underutilization of molecular diagnostic tools like PCR in the included studies, particularly for porcine cysticercosis, indicates a gap in modern surveillance techniques. Molecular confirmation is crucial for distinguishing *T. solium* from other Taenia species (Gweba *et al*., 2010).

## Conclusion

In conclusion, this meta-analysis confirms that *Taenia solium* cysticercosis/taeniasis is a widespread and significant health issue in India, with notable regional hotspots and a clear gender disparity in human infection. The strong correlation between human and porcine prevalence underscores the necessity of a “One Health” approach for effective control (Zinsstag *et al*., 2011; Okello *et al*., 2016). Future efforts should focus on targeted interventions in high-prevalence zones, promoting improved sanitation and pig husbandry, and enhancing surveillance with advanced diagnostic tools to move towards the elimination of *T. solium* as a public health problem in India.

## Data Availability

All data produced in the present study are available upon reasonable request to the authors

